# Transcriptome-driven Health-status Transversal-predictor Analysis (THTA) using the PBMC transcriptome for health, food, microbiome and disease markers for understanding the background and development of lifestyle diseases

**DOI:** 10.1101/2024.10.24.24316039

**Authors:** Tilman Todt, Inge van Bussel, Lydia Afmann, Lorraine Brennan, Diana G. Ivanova, Yoana Kiselova-Kaneva, E. Louise Thomas, Ralph Rühl

## Abstract

We developed a novel machine-learning artificial intelligence (AI) approach to predict general health and food-intake parameters named Transcriptome-driven Health-status Transversal-predictor Analysis (THTA) with relevance for diabesity markers based on a mathematics-driven and non-transcriptomic biomarker driven approach. The prediction was based on values from food consumption, dietary lipids and their bioactive metabolites, peripheral blood mononuclear cells (PBMC) mRNA-based transcriptome signatures, magnetic resonance imaging (MRI), energy metabolism measurements, microbiome analyses, and baseline clinical parameters in a cohort of 72 subjects. Our novel machine learning approach included transcriptome data from PBMCs as a “one-method” approach to predict 77 general health-status markers for broad stratification of the diabesity phenotype, which are usually necessitating measurements using 16 different methods. The PBMC transcriptome was used to determine these selected 77 basic and background health-status markers in a transversal-predictor establishment group with very high accuracy (Pearson correlations are r = 0,94 ranging from 0,88 to 0,98). These collected variables offer a valuable indication to identify which individual factor(s) are mainly targeting diabesity. Based on the “establishment group“ prediction approach a further “confirmation group” prediction approach was performed with a predictive potential for these 77 variables of r = 0,62 (ranging from 0,30 to 0,99). This “one-method” approach allows monitoring of a large number of health-status variables with relevance for diabesity simultaneously and may enable monitoring of therapeutic and preventive strategies. In summary, this novel technique based on PBMC transcriptomics from human blood offers prediction of a large range of health-related markers, which independently would be obtained in different clinical / research centres at a much higher price.

## Introduction

Diabesity is a multifaceted disease often developing from obesity towards a diabetic-obesity phenotype (Farag and Gaballa 2011; M. I. Schmidt and Duncan 2003). Many simultaneous lifestyle factors may cause the transition to diabesity like a general unhealthy lifestyle including tobacco smoking, poor sleep habits, high alcohol consumption, low physical activity, and an unhealthy diet (Altobelli et al. 2020; Mozaffarian et al. 2009), with further contributions from a hereditary component which predisposes to diabesity (Goyal et al. 2023). Many dysfunctions in this condition are related to signalling in the glucose, insulin and fatty acids pathways as well as in the inflammation / immune system (Aronoff et al. 2004; M. I. Schmidt and Duncan 2003).

Machine learning artificial intelligence (AI) has been developed to monitor, classify, and predict diabetes (Oikonomou and Khera 2023). These predictive models can be used to study risk factors of type 2 diabetes (Schwarz et al. 2009; Yu et al. 2010; Naz and Ahuja 2020; Heikes et al. 2008) or predict diabetes (Razavian et al. 2015; Zou et al. 2018; Nguyen et al. 2019). However, small sample sizes and biased selection of risk factors may lead to predictive models which perform poorly (Lai et al. 2019). Some models use the PBMC transcriptome, in addition to other analytical methods, though most research focuses on a ‘yes/no’ answer for diabetes based on a set threshold of clinical diabetes markers (Tonyan et al. 2022). Other approaches for diabesity prediction employ polymorphisms of selected diabesity-relevant genes to indicate increased diabesity prevalence (Witka et al. 2019).

The main recommendation to reduce the burden of diabesity is lifestyle change, with various strategies (Knowler et al. 2002) and methods to both monitor and optimize lifestyle choices. However, long-term lifestyle alterations can be challenging and compliance can be poor (Aladhab and Alabbood 2019), while unfortunately quick and easy drug treatment is often preferred (Garedow et al. 2023). Lifestyle interventions often include an “all-at-once-interventions-strategy”, which are perceived as quite drastic, uncomfortable and non-preferred by patients and high-risk individuals (S. K. Schmidt et al. 2020). A more detailed evaluation of which selective lifestyle intervention should be altered while having an acceptable modified lifestyle of high-risk individuals will probably result in higher compliance and better outcomes. Selected lifestyle interventions should be chosen when many variables are evaluated at once and can be offered as a “one-method” predictor approach.

A potential approach to identifying clinical and health-status markers is based on the PBMC transcriptome (Costa et al. 2021; Elliott et al. 2014). Previous metabolic phenotyping have shown that the PBMC transcriptome can reflect environmental stress (Reynés et al. 2015) or the effect of specific diets (Bouwens, Afman, and Müller 2007; Caimari et al. 2010; Oliver et al. 2013; de Mello et al. 2012). For clinical diagnostics the PBMC transcriptome can reflect various pathologies (Baine et al. 2011; Chang et al. 2013), and identify biomarkers of obesity (Caimari et al. 2010; Oliver et al. 2013; Manoel-Caetano et al. 2012). It was previously shown that transcriptomic data from PBMCs (Rodney A. Rhoades PhD 2017) reflect the current metabolic and physiological state (Burczynski and Dorner 2006; Liew et al. 2006).

The purpose of our study was to develop models as a “one-method” machine-learning approach for determining basic and background variables like food intake and general health-status markers relevant for diabesity. Our strategy does not start by using known transcriptomic biomarkers / target genes, which are known to be involved in diabesity, but instead focusing on the categorization of the human organism as a “simple” holistic mathematical matrix. We performed a full PBMC transcriptome analysis in 72 volunteers from the EU FP7 NUTRITECH cohort, providing the largest known number of variables per person of any known performed clinical intervention study with additional 10,522 analysed gene transcripts detected in PBMCs being over quantification limit. In total 780 variables were measured by 16 different techniques, including food consumption, nutritional lipids, and their bioactive metabolites, magnetic resonance imaging (MRI), energy metabolism measures, microbiome analyses, and clinical biochemistry (Rundle et al. 2023) in the Nutritech study, while from these 780 measurements 77 variables as the most diabesity-relevant markers were selected for further analysis.

## Materials and methods

The Nutritech project encompasses comprehensive phenotyping, which includes measurements related to diet, physical activity, body composition, blood biomarkers, and metabolomics, yielding 780 measurements per individual.

### Selection of diabesity-related health-status variables for further predictor analysis

To facilitate initial testing of our predictive capabilities, we had to pare this down to a more manageable number of variables, a step taken prior to any machine learning. Our goal was to select a variety of variables linked to the development of a diabesity phenotype. We also attempted to predict variables representative of the wide range of phenotyping methods employed in the Nutritech protocol, including routinely collected physical and clinical biochemical measures, to more sophisticated and expensive analyses, including MRI, microbiome analyses, and multiple lipidomic analyses for a larger range of lipids. In total 28 self-reported dietary intake measures were included relating to plant or animal-sources, sweets/snacks and alcohol (details for categorization added in supplementary material). Moreover, we included routine health assessment measures like blood pressure and physical body measurements based on MRI (fat mass and liver fat), calorimetry, and gut microbiome analyses. Further out of total 780 variables, 77 selected variables per person were chosen with major diabesity-relevance for further targeted analysis, measured by 16 different techniques, including simple methods (eg. body weight and blood pressure; methods 9 and 15). Some variables required advanced and expensive techniques, and are available only at specialized health care facilities or a handful of service laboratories worldwide (methods 2—6; 11-14).

### Dataset

Data from the EU FP7 NUTRITECH study (Rundle et al. 2023) were used to develop regression models. The EU FP7 NUTRITECH dataset includes a human cohort of 72 subjects for whom 77 different variables out of 780 available variables and additional PBMC transcriptome data of 10,522 individually expressed genes were determined as described previously (Rundle et al. 2023; Fiamoncini et al. 2018). The cohort was divided in two groups: 32 subjects in the control group and 40 subjects in the intervention group. Data was collected before intervention as well as 13 weeks later after intervention in all participants. The data from the “before intervention group” were used for the “establishment group” model development and evaluation, while the data from the “after intervention group” were used for the “confirmation group” approach.

### Modelling and evaluation

#### A) “Establishment group” (EST) for a transversal prediction analysis

A transversal prediction analysis usually describes the analysis of a larger range of variables which can be predicted based on using one broader, more complex set of variables originating from one method, which is usually a transcriptome analysis. This prediction analysis towards obtaining transversal predictors from one variable is usually done in a group of individuals at the same time in an “establishment group”.

We developed different regression models like the Random Forest (Liaw and Wiener 2002), Elastic Net (Friedman, Hastie, and Tibshirani 2010), and multiple Linear Regression (Hebbali 2024) to evaluate the best prediction model for a broader range of variables using the PBMC transcriptomic data. For each regression analysis, an individual model was developed for each of the 77 variables using data from the “establishment group”.

The following procedure was applied for each regression method: Thirty separate models were developed for each variable. For each model, the data set was randomly split into training and validation sets. The training data set included data from two-thirds of the subjects, and one-third was used for validation. Spearman correlation coefficients were used to select predictors from the training data set. Of the 10,522 transcriptomic variables, a maximum of 50 transcripts were allowed to select predictors. mRNAs with the highest absolute correlation coefficient were selected. Regression was used to create the model. Training of the model with the Elastic Net was performed with cross-validation (with ten convolutions) and optimization of the hyperparameters. No cross-validation and optimization were performed for Random Forest and multiple Linear Regression. Multiple Linear Regression models were built by the stepwise regression function in the R package ‘olsrr’ (Hebbali 2024). The model’s root mean square error and the Pearson correlation coefficient (R) were used as evaluation metrics. After training, models were evaluated using the data from the “establishment group”. Performance of the models on the training data and the whole dataset were determined using the Pearson correlation coefficient. Data analyses and model development were performed in R (version 4.2.2) and the R package ‘caret’ (version 6.0-94) (Kuhn 2008). Random forest models in the R package ‘randomForest’ (version 4.7.1.1) (Liaw and Wiener 2002), Elastic net models applied the R package ‘glmnet’ (version 4.1.8) (Friedman, Hastie, and Tibshirani 2010) and multiple linear regression in the R package ‘olsrr’ (version 0.6.0) (Hebbali 2024). Visualisation of the results was done with the R package ‘ggplot2’ (version 4.1.8) (Wickham 2016).

#### B) Applying predictors from the “establishment group” (EST) for a “confirmation group” prediction (CON) analysis

After developing the regression models in the “establishment group” (EST), the models for predicting the outcome were applied to data of a group analysed not at the same time point, named here the “confirmation group”. Predictive performance was measured using the root mean square error and the Pearson correlation coefficient. Of the 30 models established for each variable, the model with the best performance on the “confirmation group” was selected as the final model. Predictive performance of the final models was visualized by comparing the observed values with predicted values and by comparing the overall predictive performance between the three regression methods of Random Forest, Elastic Net, and multiple Linear Regression models.

## Results

Predictive models are based on machine-learning offer an alternative to existing methods for determining variables with relevance for general health, food intake and targeted in our case for diabesity. One application of this transversal-prediction model is for monitoring individual health-status variables with a novel methodology using one blood sample, followed by a simple highly standardized transcriptomic analysis and a mathematical evaluation instead of using several different complex methodologies.

The first step was the transfer predictor analysis in the “establishment group” starting from the PBMC-transcriptome with 10,522 analysed mRNA transcripts detected, which were analysed being over the quantification limit (van Bussel et al. 2019), using 77 out of 780 variables including food intake and health-related variables for 72 volunteers.

Three different mathematical methods for prediction of 77 variables in the “establishment group” approach were used starting with the Radom Forest-based model (RF) resulting in an average of r = 0,94 ranging from 0,88 to 0,98. For prediction of the food diary data (Table 1, variables 1-28) in the transversal prediction approach predictor correlations were in the range of r = 0,92 - 0,97 (EST-RF) offering the first indirect calculation of food diary data. Especially summarising data like animal-based food (r = 0,94), plant-based food (r = 0,94), snacks / sweets (r = 0,96), general healthy food (r = 95) and general non-healthy food (r = 0,97), offer a precise predictor calculation (Figure 1). These predictor correlations are highly relevant to evaluate newly but also already existing large scale databases with PBMC transcriptomic data included but without additional food diary data collection.

**Figure 1.**
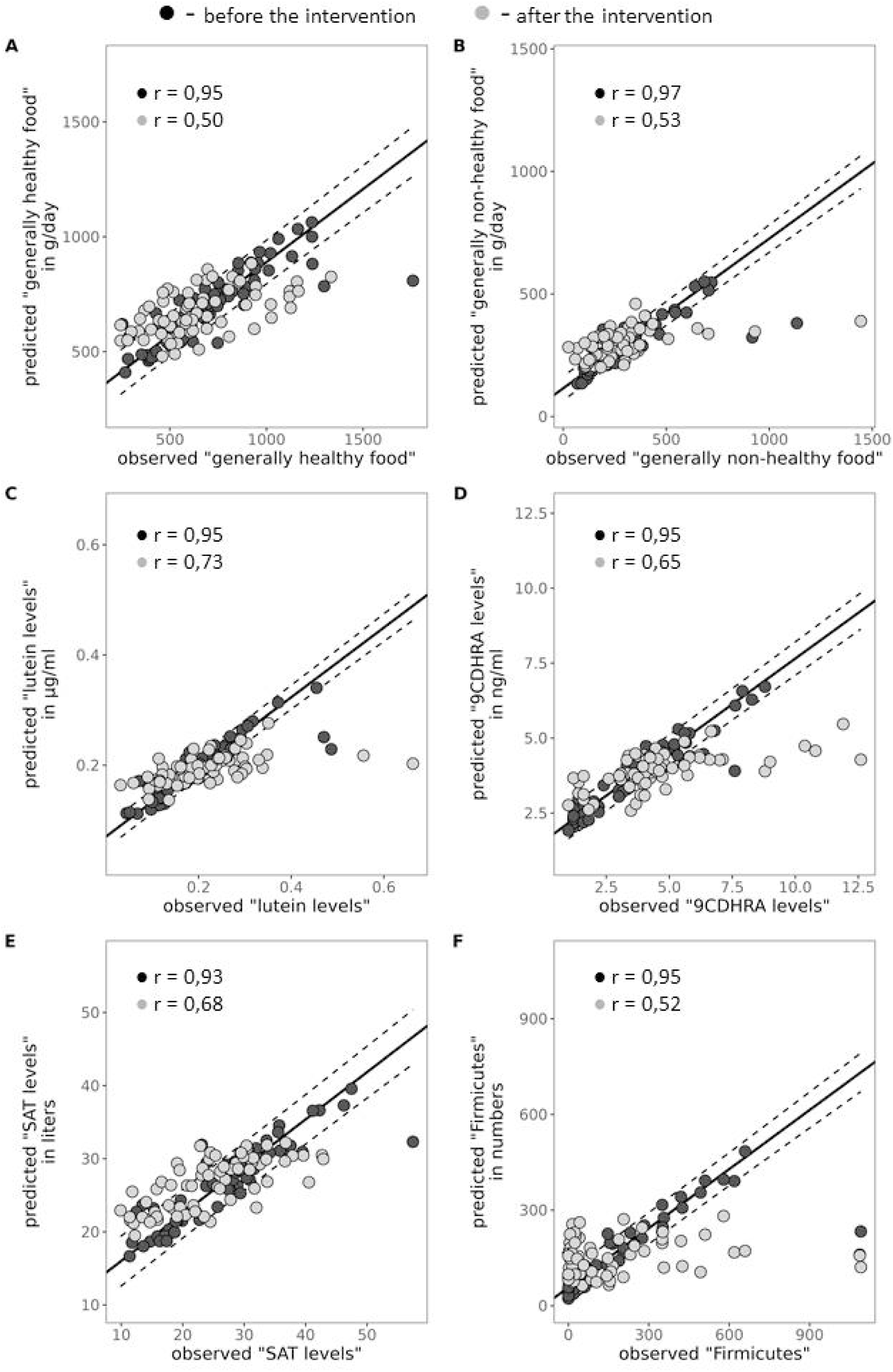
Comparison of observed and predicted values of dependent variables. For eight regression models the observed values of the dependent variable is compared to the model predicted values. Models were developed from 72 PBMC original transcriptomics using Random Forest for regression. The regression line is calculated from the observed and predicted values from the training data. The 95% confidence interval of the prediction model is indicated by the dashed lines. The dots represent the observed and predicted values of the measurements before intervention (black) and after intervention (grey). The correlation coefficient R and the percentage of measurements with the 95% confidence interval (CI) is provided. The following dependent variables are reported: (A) general healthy food; (B) general non-healthy food; (C) serum level of 9-cis-13,14-dihydroretinoic acid (9CDHRA); (D) serum level of lutein; (E) SAT levels; (F) Firmicutes numbers.

**Table 1:**
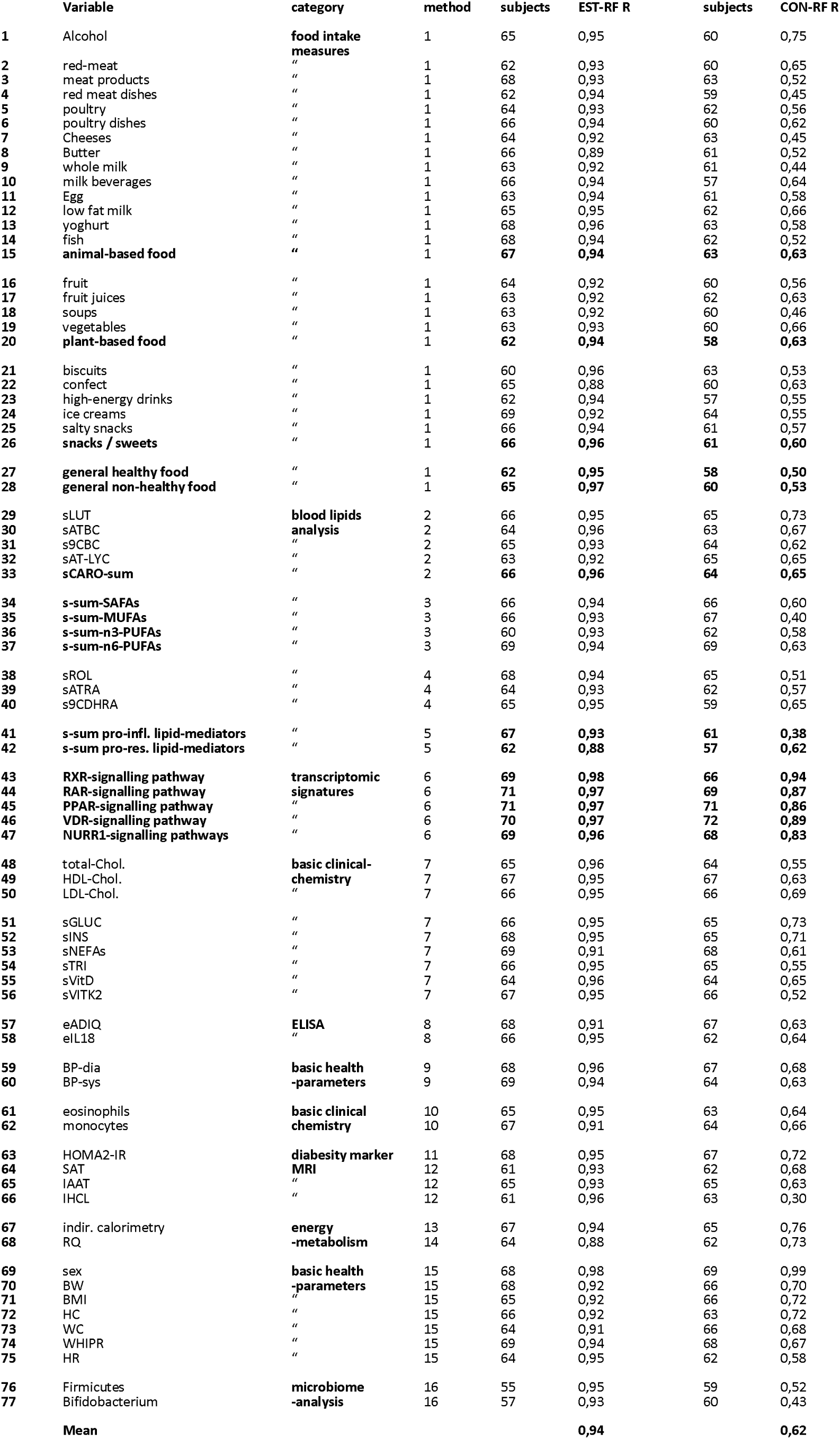
Prediction of variables using Random Forest-based model. Variables were predicted in the “establishment group” (EST) for a transversal prediction starting from the PBMC transcriptome towards the 77 listed variables using the Random Forest-based (RF) regression models and in addition a “confirmation group” prediction (CON) was performed using the same methodology in the same group of volunteers after intervention using the 72 PBMC transcriptomic data of these volunteers. For each variable the following is reported: the number of individual transcriptomics included in the datasets, the number of mRNAs the model is based on, the Pearson correlation coefficient (R) between observed and predicted values, and the experimental method used to determine the variables.

The second sets of parameters to be determined were blood lipids (Table 1, variables 29 – 42). The models performed well for variables like the sum of carotenoids (sCARO-sum, variable 33) r = 0,96, sum of saturated fatty acids (s-sum SAFAs) r = 0,94, sum of monounsaturated fatty acids (s-sum MAFAs) r = 0,93, sum of n3-polyunsaturated fatty acids (s-sum n3-PUFAs) r = 0,93 and sum of n6-polyunsaturated fatty acids (s-sum n6-PUFAs) r = 0,94, sum of pro-inflammatory lipid-mediators (s-sum pro-infl.-lipid mediators) r = 0,93 and sum of pro-resolving lipid-mediators (s-sum pro-res.-lipid mediators) r = 0,88 are precise predictor calculations of an indirect quantification of the individual lipid levels. These predictor calculations are highly relevant to evaluate newly but also already existing large scale databases with PBMC transcriptomic included to further predict these specific lipids in individuals which are usually are just available using expensive and highly laborious blood lipid analyses.

The third set of new variables (Table 1, variables 43 – 47), which indicates transcriptome-based nuclear hormone selective signalling pathways including RXR- (r = 0,98), RAR- (r = 0,97), PPAR- (r = 0,97), VDR- (r = 0,97) and NURR1 / NR4A2-signalling pathways (r = 0,96), indicating which nuclear hormone signalling pathway is currently relatively up- or down-regulated.

All health variables including basic clinical chemistry-, basic health-, body composition, and microbiome parameters (Table 1, variables 48 – 77) commonly used to determine individual health-status relevant for diabesity, were analysed. Of special interest were body composition parameters from MRI analyses including subcutaneous adipose tissue (SAT, r = 0,93), intra-abdominal adipose tissue (IAAT, r = 0,93), and intrahepatic cellular lipid (indicating liver fat, IHCL, r = 0,96). The model was also able to predict microbiome analysis including Firmicutes (r = 0,95) and Bifidobacterium (r = 0,93).

Other transfer predictor regression models like the Elastic Net (EN) and the Linear Regression model (LR) were tested. Predictor correlations suggested that the Random Forest model performed best with an average of r = 0,94, followed by the Elastic Net model r = 0,80 and the Linear Regression model r = 0,77 (table 2, supplementary tables 2 and 3 and figure 2). Applying all the methodologies and calculating a “best of” predictor analysis offer correlation coefficients of r = 0,94. Thus, the Random Forest model always offered the best values for the “establishment group”.

**Figure 2.**
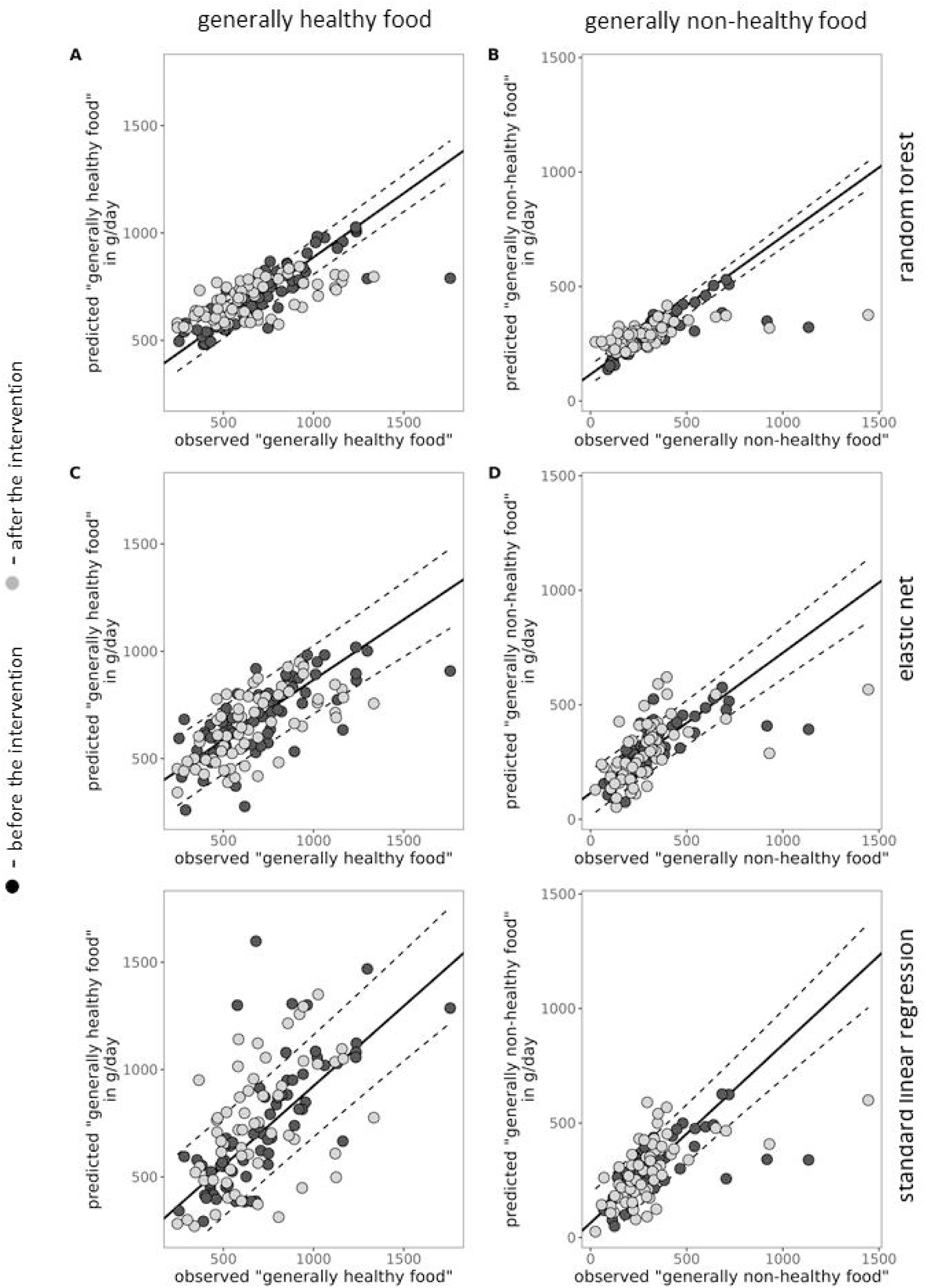
Comparison of the performance of regression methods. For the variable ‘general healthy food’ and ‘general non-healthy food’ regression models were generated using Random Forest (A and B), Elastic Net (C and D), and a standard linear regression method (E and F). Models were developed from 72 PBMC transcriptomic data. The regression line is calculated from the observed and predicted values from the training data. The 95% confidence interval (CI) of the prediction model is indicated by the dashed lines. The dots represent the observed and predicted values of the measurements before intervention (black) and after intervention (GREY). The correlation coefficient R and the percentage of measurements with the 95% CI is provided.

**Table 2:** Prediction of variables using Random Forest-based (RF) regression model, the Elastic Net (ER) regression model or the Liner Regression (LR) model and calculating the “best of” (BO) three methods. Summary of the transversal prediction analysis in the “establishment group” (EST) using 77 variables starting from the PBMC transcriptome using the Random Forest-based (RF) regression model, the Elastic Net (ER) regression model or the Liner Regression (LR) model. A further prediction in a “confirmation group” prediction (CON) was performed for the same 77 variables using the same methodology in the same group of volunteers after intervention using the 72 PBMC transcriptomic data of these volunteers. For each variable the following is reported: the number of individual transcriptomics included in the datasets, the number of mRNAs the model is based on, the Pearson correlation coefficient (R) between observed and predicted values, the R-squared (R2) of the model, and the experimental method used to determine the variables. For abbreviations check table 1.

|  | variable | EST-RF R | EST-EN R | EST-LR R | EST-BO R | CON-RF R | CON-EN R | CON-LR R | CON-BO R |
| --- | --- | --- | --- | --- | --- | --- | --- | --- | --- |
| 1 | alcohol | 0,95 | 0,75 | 0,78 | 0,95 | 0,75 | 0,70 | 0,57 | 0,75 |
| 2 | red-meat | 0,93 | 0,79 | 0,77 | 0,93 | 0,65 | 0,64 | 0,62 | 0,65 |
| 3 | meat products | 0,93 | 0,84 | 0,79 | 0,93 | 0,52 | 0,45 | 0,50 | 0,52 |
| 4 | red meat dishes | 0,94 | 0,75 | 0,73 | 0,94 | 0,45 | 0,40 | 0,32 | 0,45 |
| 5 | poultry | 0,93 | 0,82 | 0,84 | 0,93 | 0,56 | 0,58 | 0,44 | 0,58 |
| 6 | poultry dishes | 0,94 | 0,89 | 0,81 | 0,94 | 0,62 | 0,66 | 0,59 | 0,66 |
| 7 | cheeses | 0,92 | 0,67 | 0,63 | 0,92 | 0,45 | 0,34 | 0,29 | 0,45 |
| 8 | butter | 0,89 | 0,73 | 0,51 | 0,89 | 0,52 | 0,36 | 0,41 | 0,52 |
| 9 | whole milk | 0,92 | 0,68 | 0,72 | 0,92 | 0,44 | 0,47 | 0,33 | 0,47 |
| 10 | milk beverages | 0,94 | 0,90 | 0,83 | 0,94 | 0,64 | 0,56 | 0,55 | 0,64 |
| 11 | egg | 0,94 | 0,72 | 0,75 | 0,94 | 0,58 | 0,62 | 0,56 | 0,62 |
| 12 | low fat milk | 0,95 | 0,75 | 0,72 | 0,95 | 0,66 | 0,74 | 0,66 | 0,74 |
| 13 | yoghurt | 0,96 | 0,82 | 0,77 | 0,96 | 0,58 | 0,68 | 0,60 | 0,68 |
| 14 | fish | 0,94 | 0,83 | 0,77 | 0,94 | 0,52 | 0,61 | 0,45 | 0,61 |
| 15 | animal-based food | 0,94 | 0,81 | 0,74 | 0,94 | 0,63 | 0,60 | 0,50 | 0,63 |
| 16 | fruit | 0,92 | 0,68 | 0,68 | 0,92 | 0,56 | 0,59 | 0,39 | 0,59 |
| 17 | fruit juices | 0,92 | 0,75 | 0,72 | 0,92 | 0,63 | 0,48 | 0,41 | 0,63 |
| 18 | soups | 0,92 | 0,76 | 0,69 | 0,92 | 0,46 | 0,39 | 0,41 | 0,46 |
| 19 | vegetables | 0,93 | 0,80 | 0,86 | 0,93 | 0,66 | 0,66 | 0,54 | 0,66 |
| 20 | plant-based food | 0,94 | 0,80 | 0,80 | 0,94 | 0,63 | 0,62 | 0,54 | 0,63 |
| 21 | biscuits | 0,96 | 0,81 | 0,79 | 0,96 | 0,53 | 0,51 | 0,49 | 0,53 |
| 22 | confect | 0,88 | 0,70 | 0,73 | 0,88 | 0,63 | 0,59 | 0,58 | 0,63 |
| 23 | high-energy drinks | 0,94 | 0,74 | 0,74 | 0,94 | 0,55 | 0,64 | 0,55 | 0,64 |
| 24 | ice creams | 0,92 | 0,72 | 0,69 | 0,92 | 0,55 | 0,51 | 0,51 | 0,55 |
| 25 | salty snacks | 0,94 | 0,76 | 0,72 | 0,94 | 0,57 | 0,59 | 0,51 | 0,59 |
| 26 | snacks / sweets | 0,96 | 0,86 | 0,81 | 0,96 | 0,60 | 0,69 | 0,61 | 0,69 |
| 27 | general healthy food | 0,95 | 0,85 | 0,86 | 0,95 | 0,50 | 0,51 | 0,31 | 0,51 |
| 28 | general non-healthy food | 0,97 | 0,88 | 0,83 | 0,97 | 0,53 | 0,63 | 0,59 | 0,63 |
| 29 | sLUT | 0,95 | 0,80 | 0,74 | 0,95 | 0,73 | 0,78 | 0,65 | 0,78 |
| 30 | sATBC | 0,96 | 0,73 | 0,74 | 0,96 | 0,67 | 0,74 | 0,69 | 0,74 |
| 31 | s9CBC | 0,93 | 0,88 | 0,77 | 0,93 | 0,62 | 0,62 | 0,63 | 0,63 |
| 32 | sAT-LYC | 0,92 | 0,85 | 0,87 | 0,92 | 0,65 | 0,67 | 0,56 | 0,67 |
| 33 | sCARO-sum | 0,96 | 0,88 | 0,81 | 0,96 | 0,65 | 0,66 | 0,58 | 0,66 |
| 34 | s-sum-SAFAs | 0,94 | 0,81 | 0,68 | 0,94 | 0,60 | 0,62 | 0,63 | 0,63 |
| 35 | s-sum-MUFAs | 0,93 | 0,73 | 0,62 | 0,93 | 0,40 | 0,50 | 0,43 | 0,50 |
| 36 | s-sum-n3-PUFAs | 0,93 | 0,89 | 0,83 | 0,93 | 0,58 | 0,72 | 0,60 | 0,72 |
| 37 | s-sum-n6-PUFAs | 0,94 | 0,84 | 0,83 | 0,94 | 0,63 | 0,70 | 0,63 | 0,70 |
| 38 | sROL | 0,94 | 0,78 | 0,71 | 0,94 | 0,51 | 0,48 | 0,44 | 0,51 |
| 39 | sATRA | 0,93 | 0,78 | 0,72 | 0,93 | 0,57 | 0,38 | 0,45 | 0,57 |
| 40 | s9CDHRA | 0,95 | 0,87 | 0,83 | 0,95 | 0,65 | 0,72 | 0,71 | 0,72 |
| 41 | s-sum pro-infl. lipid-mediators | 0,93 | 0,75 | 0,75 | 0,93 | 0,38 | 0,41 | 0,20 | 0,41 |
| 42 | s-sum pro-res. lipid-mediators | 0,88 | 0,75 | 0,71 | 0,88 | 0,62 | 0,64 | 0,49 | 0,64 |
| 43 | RXR-signalling pathway | 0,98 | 0,96 | 0,96 | 0,98 | 0,94 | 0,96 | 0,96 | 0,96 |
| 44 | RAR-signalling pathway | 0,97 | 0,96 | 0,96 | 0,97 | 0,87 | 0,95 | 0,95 | 0,95 |
| 45 | PPAR-signalling pathway | 0,97 | 0,91 | 0,91 | 0,97 | 0,86 | 0,87 | 0,85 | 0,87 |
| 46 | VDR-signalling pathway | 0,97 | 0,94 | 0,92 | 0,97 | 0,89 | 0,91 | 0,88 | 0,91 |
| 47 | NURR1-signalling pathways | 0,96 | 0,94 | 0,94 | 0,96 | 0,83 | 0,90 | 0,89 | 0,90 |
| 48 | total-Chol. | 0,96 | 0,76 | 0,74 | 0,96 | 0,55 | 0,65 | 0,59 | 0,65 |
| 49 | HDL-Chol. | 0,95 | 0,80 | 0,76 | 0,95 | 0,63 | 0,68 | 0,63 | 0,68 |
| 50 | LDL-Chol. | 0,95 | 0,86 | 0,82 | 0,95 | 0,69 | 0,71 | 0,63 | 0,71 |
| 51 | sGLUC | 0,95 | 0,83 | 0,76 | 0,95 | 0,73 | 0,76 | 0,59 | 0,76 |
| 52 | sINS | 0,95 | 0,82 | 0,75 | 0,95 | 0,71 | 0,69 | 0,63 | 0,71 |
| 53 | sNEFAs | 0,91 | 0,67 | 0,59 | 0,91 | 0,61 | 0,61 | 0,57 | 0,61 |
| 54 | sTRI | 0,95 | 0,78 | 0,62 | 0,95 | 0,55 | 0,58 | 0,62 | 0,62 |
| 55 | sVITD | 0,96 | 0,82 | 0,74 | 0,96 | 0,65 | 0,70 | 0,69 | 0,70 |
| 56 | sVITK2 | 0,95 | 0,82 | 0,82 | 0,95 | 0,52 | 0,60 | 0,53 | 0,60 |
| 57 | eADIQ | 0,91 | 0,85 | 0,77 | 0,91 | 0,63 | 0,70 | 0,68 | 0,70 |
| 58 | eIL18 | 0,95 | 0,85 | 0,72 | 0,95 | 0,64 | 0,51 | 0,56 | 0,64 |
| 59 | BP-dia | 0,96 | 0,83 | 0,76 | 0,96 | 0,68 | 0,70 | 0,67 | 0,70 |
| 60 | BP-sys | 0,94 | 0,76 | 0,76 | 0,94 | 0,63 | 0,63 | 0,53 | 0,63 |
| 61 | eosinophils | 0,95 | 0,85 | 0,82 | 0,95 | 0,64 | 0,61 | 0,45 | 0,64 |
| 62 | monocytes | 0,91 | 0,70 | 0,68 | 0,91 | 0,66 | 0,59 | 0,47 | 0,66 |
| 63 | HOMA2-IR | 0,95 | 0,79 | 0,77 | 0,95 | 0,72 | 0,72 | 0,64 | 0,72 |
| 64 | SAT | 0,93 | 0,81 | 0,78 | 0,93 | 0,68 | 0,73 | 0,69 | 0,73 |
| 65 | IAAT | 0,93 | 0,77 | 0,76 | 0,93 | 0,63 | 0,69 | 0,70 | 0,70 |
| 66 | IHCL | 0,96 | 0,82 | 0,83 | 0,96 | 0,30 | 0,28 | 0,15 | 0,30 |
| 67 | indir. calorimetry | 0,94 | 0,87 | 0,91 | 0,94 | 0,76 | 0,69 | 0,64 | 0,76 |
| 68 | RQ | 0,88 | 0,69 | 0,67 | 0,88 | 0,73 | 0,67 | 0,44 | 0,73 |
| 69 | sex | 0,98 | 0,94 | 0,93 | 0,98 | 0,99 | 0,97 | 0,98 | 0,99 |
| 70 | BW | 0,92 | 0,77 | 0,74 | 0,92 | 0,70 | 0,75 | 0,73 | 0,75 |
| 71 | BMI | 0,92 | 0,69 | 0,66 | 0,92 | 0,72 | 0,74 | 0,61 | 0,74 |
| 72 | HC | 0,92 | 0,80 | 0,76 | 0,92 | 0,72 | 0,69 | 0,53 | 0,72 |
| 73 | WC | 0,91 | 0,81 | 0,78 | 0,91 | 0,68 | 0,74 | 0,71 | 0,74 |
| 74 | WHIPR | 0,94 | 0,85 | 0,81 | 0,94 | 0,67 | 0,65 | 0,58 | 0,67 |
| 75 | HR | 0,95 | 0,78 | 0,77 | 0,95 | 0,58 | 0,54 | 0,52 | 0,58 |
| 76 | Firmicutes | 0,95 | 0,92 | 0,89 | 0,95 | 0,52 | 0,55 | 0,49 | 0,55 |
| 77 | Bifidobacterium | 0,93 | 0,68 | 0,51 | 0,93 | 0,43 | 0,37 | 0,39 | 0,43 |
|  | Mean | 0,94 | 0,80 | 0,77 | 0,94 | 0,62 | 0,63 | 0,57 | 0,65 |

Further, we evaluated the robustness of this methodology by the “establishment group” predictor calculations for variables all determined “at the same day” with variables obtained weeks later in the same person; half of the group maintained their initial lifestyle, and the second group underwent a lifestyle intervention with increased physical activity, an energy-restricted and more balanced diet (Rundle et al. 2023; Fiamoncini et al. 2018). In this second group, the “confirmation group” prediction (CON) approach, we did not dissect these two groups for further calculations but this will be done under future evaluation of the 77 selected health-status parameters for a detailed evaluation of success of a lifestyle change on diabesity.

In these confirmation predictions, the Elastic Net model (r = 0,63) provided the best predictive potential, followed by the Random Forest model (r = 0,62) and by the Linear Regression model r = 0,57 (Table 2 and supplementary tables 2 and 3). Applying all actual methodologies and calculating a “best of” predictor analysis offer values of r = 0,65. Consequently, the “best of” predictor calculation offered the best values for the confirmation group mainly based on the Random Forest and Elastic Net models (Figure 3).

**Figure 3.**
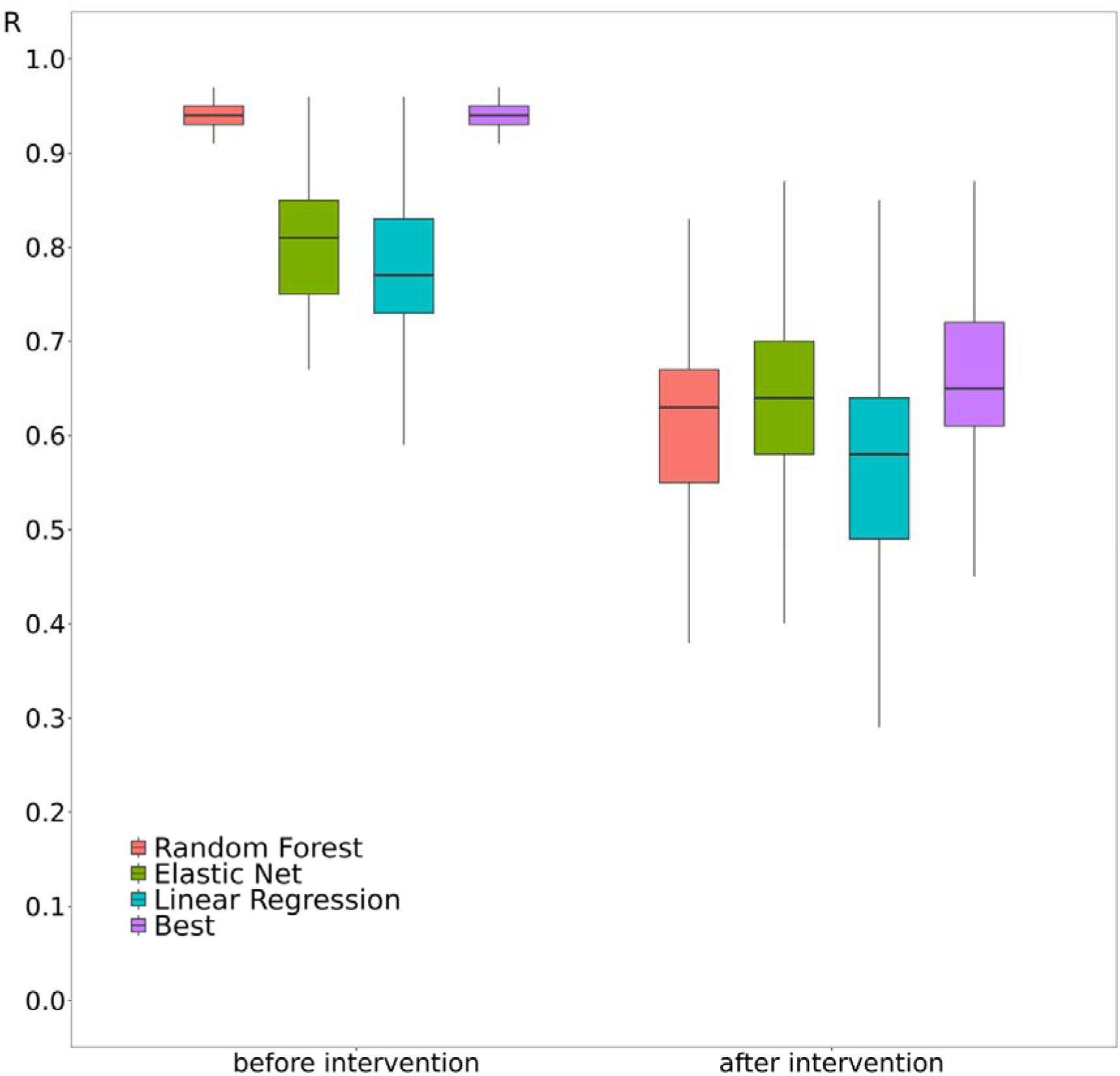
Summarizing figure of the establishment transversal-prediction analysis (on the left side) ranging from the Random Forest, Elastic Net and Linear Regression model prediction analyses as well as “best of” (best) summarising prediction models in addition to confirmation predictions (at the right side) based on the Random Forest, Elastic Net and Linear Regression model prediction analysis as well as “best of” (best) summarising prediction model.

## Discussion

Machine-learning approaches for health-status monitoring relevant for diabesity (Agliata et al. 2023; Wee et al. 2024) have been developed recently, and many of the existing machine-learning models use common biomarkers / risk factors and variables mainly associated with diabetes (Okada et al. 2022). These prediction models are mainly targeting single variables and are based on preselected known biomarkers (Jangili et al. 2023). Until now, no broader health-status marker array analyses have been performed with a focus on diabesity.

We used data from the EU FP7-NUTRITECH consortium including 72 individuals with 780 analysed individual clinical and lifestyle parameters and additional 10,522 parameters from PBMC transcriptome analyses. For our transfer-predictor analysis we used these 10,522 transcriptomic variables from mRNA transcripts only based on mathematical-relevance and not based on known biomarker functions, which is a new approach. This novel design simply applies data from the PBMC transcriptomics as a “simple” holistic and complex mathematical matrix (Figure 4).

**Figure 4.**
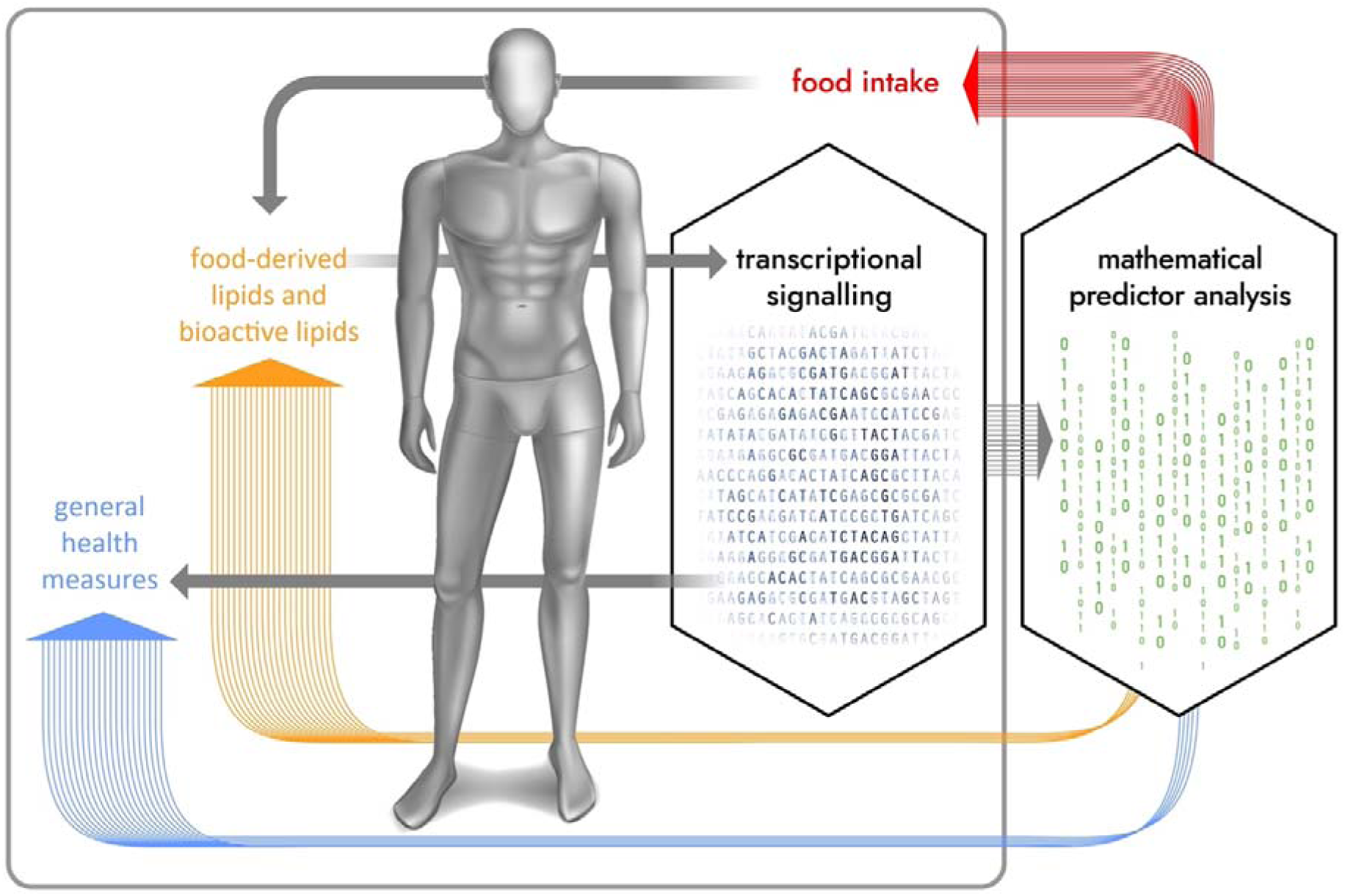
Schematic model of our transversal-predictor analysis

These data from PBMC can easily be obtained from blood samples. A previous study has shown that individual PBMC transcriptomes can reflect obesity-related inflammation (Bories et al. 2012) and lipid metabolism in obesity-associated organs (Konieczna et al. 2015). Our approach determined that our “single method” predictor analysis could predict many individual lifestyle parameters such as food intake, basal and advanced health-status markers that were measured in the NUTRITECH cohort. In addition, as it is a “clean” mathematical methodology where we just focussed on the 77 most relevant health–status variables from a total of 780 parameters measured in the NUTRITECH cohort at one time point. We also selectively tested the possibility that alternative parameters of these 780 parameters can be calculated with similar accuracy using our transversal-predictor analysis (data not shown).

Diabetes mellitus is a multifactorial chronic disease, which can be triggered by many genetic and/or environmental factors (Sun, Yu, and Hu 2014; Kaul and Ali 2016). The number of adults living with diabetes globally has almost quadrupled since 1980 to 422 million and is projected to increase to 693 million by 2045 (Cho et al. 2018). Lifestyle interventions can be very effective in preventing and treating diabetes (Knowler et al. 2002), whereas they often are not well tolerated by participants (S. K. Schmidt et al. 2020). The inclusion of multiple diabetes-related variables may allow the development of targeted, more selective, and potentially more acceptable lifestyle interventions (Zhang et al. 2020). A cost-effective method would allow monitoring to be performed more frequently. In our novel machine-learning approach, we have developed regression models that can be used as low-cost models with good predictive power for continuous monitoring and evaluation of lifestyle interventions.

Furthermore, our approach provides a novel methodology for analysing and monitoring personal health status. Most current approaches use predictive models to distinguish between groups of individuals (e. g. healthy versus diabetic) through classification and use of common risk factors or health-related variables for model development. Our approach does not require the separation of individuals beforehand and uses PBMC transcriptome and a mathematical approach to evaluate many health-status markers concerning individual metabolic and physiological status.

The ability to robustly predict dietary intake, without necessitating the collection and analysis of food intake diaries is an exciting advancement. In the NUTRITECH studies these food dairy data can now easily be used for further correlations with clinical chemistry and this transversal-prediction approach. Especially summarised food groups like healthy or unhealthy dietary patterns, snacks / sweets, animal-based foods, or plant-based foods can be precisely predicted with high accuracy (r = 0,94 – 0,97) (Table 2).

Our methodology offers a straightforward and cost-effective approach to predicting and monitoring 77 most relevant health-status markers. In our analysis, the predictive models demonstrated excellent accuracy within the “establishment group” but showed moderate accuracy in the “confirmation group”. The predictive power in the “confirmation group” is influenced mainly by the availability of parameters with a larger distribution pattern and reduces accuracy.

Larger cohort studies are needed to further validate and improve the robustness of our approach. A potential limitation of these studies is their typically more shallow approach to phenotyping, which may lack the comprehensive range of parameters available in studies such as NUTRITECH (Rundle et al. 2023; Fiamoncini et al. 2018). This difference could impact the depth and accuracy of this transversal-predictor analysis when applied to broader datasets. However, these studies would enable the inclusion of additional parameters, such as sex, hormone levels, medication use, disease status, age, and ethnicity, which could enhance the models’ predictive power. They would also support the development of specialized models for specific subgroups, including pregnant women, children, athletes, individuals with unique genetic profiles, older adults, and those with specific diseases. These expansions would optimize the methodology, increase predictive accuracy, and broaden its applicability across diverse populations, allowing for more personalized health monitoring and prediction.

In summary, we have developed a novel, “one-method” machine-learning approach for transversal-prediction in an “establishment group”, termed “Transcriptome-driven Health-status Transversal-predictor Analysis” (THAP). This method was further validated in a “confirmation group” consisting of 72 individuals, leveraging PBMC transcriptome data and mathematical processing to predict 77 of the most relevant health-status variables from an original set of 780 variables. This method introduces a novel predictive model that offers a cost-effective approach for monitoring health status markers, aimed at optimizing lifestyle choices and understanding their impact on the development and progression of health conditions, particularly diabesity. Our approach demonstrates that leveraging the individual PBMC transcriptome as a holistic mathematical matrix serves as a robust source for personalized health predictions. This model may also represent a simple yet innovative approach to clinical diagnosis and the management of dietary interventions and pharmacotherapy.

## Data Availability

All data produced in the present study are available upon reasonable request to the authors

## Abbreviations

For food intake measures please check the materials and methods

s: serum
e: ELISA
adj.: adjusted
sLUT: serum lutein
sATBC: serum all-trans-β-carotene
s9CBC: serum 9-cis-β-carotene
sATLYC: serum all-trans-lycopene
CAROsum: sum of total carotenoids in serum
SAFA: saturated fatty acids
MUFA: mono-unsaturated fatty acids
PUFAs: polyunsaturated fatty acids
sROL: retinol
sATRA: serum all-trans- retinoic acid
s9CDHRA: serum 9-cis 13,14-dihydroretinoic acid
pro-inf.: proinflammatory
pro-res.: pro- resolving
RXR: retinoid X receptor
RAR: retinoic acid receptor
PPAR: peroxisome proliferator-activated receptor
LXR: liver X receptor
VDR: vitamin D receptor
NURR1: nuclear receptor related 1 protein/ NR4A2- receptor
Chol: cholesterol
HDL: high density lipoprotein
LDL: low density lipoprotein
GLUC: glucose
INS: insulin
NEFAS: non-esterified fatty acids
TRI: triglycerides
VitD: 25-Hydroxyvitamin D3
VitK2: vitamin K2
ADIQ: adiponectin
IL18: interleukin 18
BP-dia: diastolic blood pressure
BP-sys: systolic blood pressure
HOMA2-IR: homeostasis model assessment index
SAT: subcutaneous adipose tissue
IAAT: intraabdominal adipose tissue
IHCL: intrahepatic cellular lipid index
indir calorimetry: indirect calorimetry
RQ: respiratory coefficient
BW: body weight
BMI: body weight index
HC: hip circumcision
WC: waist circumcision
WHIPR: waist hip ratio
HR: heart rate.

