## Supplemental material for "Transcriptome-driven Health-status Transversal-predictor Analysis (THTA) using the PBMC transcriptome for health, food, microbiome and disease markers for understanding the background and development of lifestyle diseases"

***Supplementary material***

**Recruitment and study population**

Research Ethics was granted by the West London Ethic Committee (12/LO/0139) and is registered on the clinical trials website (ClinicalTrials.gov NCT01684917). Inclusion and exclusion criteria are shown in Supplemental Table 1.

**Screening visit**

Following initial phone screening, volunteers attended a health-screening visit at the research facility. The assessment included height, weight, body fat percentage using bioelectric impedance, blood pressure, a 12 lead electrocardiography (ECG), and blood sampling to measure glucose, insulin, full blood count (FBC) and lipids. Details of medical history, medications and lifestyle were collected; current dieters were not included in the study. Volunteers currently on medications not interfering with metabolism (otherwise excluded) and dietary supplements were asked to continue the use throughout the study.

**Anthropometry**

Body weight (kg) was measured using a Seca scale (Vogel & Halke Hamburg, D). Height (m) was recorded using a stadiometer (Invicta Plastics Ltd., Leicester, UK). Waist and hip circumferences (cm) were measured by an experienced observer. Waist circumference was measured at the (WHO recommended) midpoint between the distal border of the lowest rib and the superior border of the iliac crest ((1)). Body mass index (BMI kg/m^2^) and waist to hip ratio (WHR) were calculated from these parameters.

**Classical biochemistry**

Total cholesterol, HDL-cholesterol, LDL-cholesterol, and triglycerides were measured using standard methodologies performed by the Department of Biochemistry, Molecular Medicine and Nutrigenomics of the Medical University of Varna, Bulgaria using an Olympus AU400 analyser.

**Measurement of biomarkers**

Multiple biomarkers including glucose, insulin, nonesterified fatty acids (NEFA), interleukin (IL)18, leptin (LEP) and adiponectin (ADIQ) were quantified in plasma. Plasma concentration of glucose was measured using Abbott Architect ci8200 analyser (Abbott Diagnostics, USA) at Department of Biochemistry, Hammersmith Hospital. Serum insulin levels were measured using the Millipore Human Insulin Specific RIA HI-14k kit (Millipore Corporation, Billerica, USA) accordingly to manufacturer’s instructions.

**Magnetic resonance imaging (MRI)**

Detailed methodology of the MRI measurements to quantify total and regional adipose tissue (AT) depots, as well as fat content of the liver, pancreas, and the soleus and tibialis muscles, have been reported elsewhere (2, 3).

**Calculation of metabolic indices**

The metabolic index HOMA2_IR (*HOMA-IR = (fasting glucose x fasting insulin)* was calculated to provide insight in systemic and organ-specific changes in glucose- and insulin-related metabolism (4).

**Energy expenditure**

Resting energy expenditure was assessed by indirect calorimetry (GEM, UK) on day 1 of each assessment week in a fasted state. Participants were asked to rest for 15 min in semi-recumbent position before the measurement. Calibration and measurement were collected using a well-established method (5). Large transparent canopy was placed over the head and thorax. Once the carbon dioxide content of the air entering the chamber stabilized, participants were asked to stay still for 20 min during gas collection. Resting metabolic rate (RMR) and respiratory quotient (RQ) were obtained.

**PBMC RNA isolation and microarray processing and microarray data analysis** were performed as previously described (6).

**Food frequency analysis**

Habitual food intake was assessed in week 1 by a 7-day food diary and again in week 13 to check for compliance to the prescribed diet. Subjects were instructed to estimate portion sizes using household measures like cups and tablespoons. Brand names were also given where appropriate. Food diaries were analysed using dietary analysis software Diet Plan 6.70 software (Forestfield software Ltd, 1991-2012) for complete macro- and micronutrients profile; food portion size book ((7)) was used where food quantities were omitted from the diary. Additional categories were formed by clustering single food intake categories. The additional categories are listed below.

| **Category** | **Contains** |
| --- | --- |
| Animal-based food | red meat, meat products, red meat dishes, poultry, poultry dishes, cheese, butter, whole milk, milk beverages, egg, low fat milk, yoghurt, fish |
| Plant-based food | fruit, fruit juices, soups, vegetables |
| Snacks/ sweets | biscuit, chips, confect, high-energy drinks, ice cream, salty snacks |
| General healthy food (GHF) | fish, yoghurt, fruit, soups, vegetables |
| General non-healthy food (GNHF) | red meat, meat products, red meat dishes, snacks / sweets, |

**Retinoid analysis**

High performance liquid chromatography mass spectrometric (HPLC-MS; 2695XE separation module; Waters, Hungary)–mass spectrometry (Micromass Quattro Ultima PT; Waters, Hungary) analyses were performed under dark yellow/amber light using a previously validated protocol for the determination of all-*trans*-retinoic acid and retinol in serum and tissue samples (8) and optimized further for dihydro-retinoids (9, 10).

**Eicosanoid analysis**

HPLC-ESI-MS-MS analysis of free fatty acids and eicosanoids and docosanoids in tissue (skin, PBMCs and adipose tissue) and serum. The previously described procedure for the HPLC-ESI-MS-MS method was followed with minor modifications (11, 12).

*Sample preparation*. The analytical sample preparation procedure is based on an established method for retinoid quantification 22. In summary, to 50 mg of WAT (if less than 50 mg of WAT biopsy was present, water was added to yield 50 mg of sample weight), then 150 μl acetonitrile was added and the biopsies were cut with scissors in small pieces on ice. These mixtures were shaken for 3 min, the precipitated protein was centrifuged at 13.000 rpm, 4°C for 6 min, 130 μL of the resulting supernatant was spiked with 10 μl isotope supernatant mix, evaporated in Eppendorf reaction vials with an Eppendorf concentrator at 30°C for ~60 min until the sample volume was ~10 µl. The Eppendorf concentrator was vented with argon to prevent degradation of eicosanoids and docosanoids. The dried extract was resuspended with approximately 25 μl of HPLC solvent A [64.3% water (water, Chromasolv Plus from Sigma-Aldrich, H), and 35.5% acetonitrile (Merck KGaA, D), and 0.2% formic acid (Fluka, H)] to yield 35 μl, then vortexed (15 sec), shaken (3 min) and transferred into micro injection inserts vials (Waters, H). These glass vials with the 35 μl extract were transferred into brown screw top vials with PTFE/ silicone septa sample (Waters, H) and placed in the pre-cooled (15°C) autosampler of the Waters 2695XE separation module.

*Chromatographic system*. The HPLC system consisted of a Waters 2695XE separation module (Waters, H) including a gradient pump, autosampler, degasser and a heated column compartment. A MS-MS detector with an ESI ionizing option was used (Micromass Quattro Ultima PT from Waters, UK) as a detector. The system was controlled via the MassLynx software (Waters, H).

*HPLC conditions*. The eluents were degassed in the Waters 2695XE separation module prior to mixing, then passed through an in-line filter (1-2 μm; Knauer, D) before reaching the analytical column (LiChroCART, 125 X 2 mm; Superspher 100, RP-18, endcapped) from Merck KgaA (D) embedded in the column compartment. A multilinear gradient was formed from solvent A (see above) and solvent B (methanol; Merck KGaA D). The gradient included the following steps: 0.0 min 20% B; 3.0 min 20 % B; 5.0 min 60 % B; 15.0 min 100 % B; 15.9 min 100 % B and 16.0 min 5 % B. The flow rate was adjusted to 0.4 mL/min and the column was heated to 40°C. From the same biological extract, 10 μL for each HPLC analysis was used. This step was performed twice using the same HPLC conditions and two different MS-MS analysis options for better resolution and quantification of the various analytes.

MS options; The Micromass Quattro Ultima PT was controlled via the MassLynx software. Argon with an inlet pressure of 0.8 bar was used. ESI (electro spray ionization source, Waters, H) was vented by nitrogen continuously produced by the nitrogen generator (Peak Scientific NM30 Nitrogen generator) including compressor (Waters, H) with the inlet flow set at 3.6 e-3 mbar.

*Multiple reaction monitoring settings*. ESI, with a negative ESI – setting, was performed with the HPLC eluent following the ion source temperature of 85°C. The desolvation gas flow was 780 L/h, the desolvation temperature was 400°C, the cone gas flow was 10 L/h, the capillary current was 3 μA, and the cone voltage was 50 V. Aperture voltage was set at 0 V and the RF lens voltage was set at 35 V (for 1) and 0.2 V (for 2). The analyzer settings were LM1 resolution 14.5; HM1 resolution 14.5; ion energy 1 0.7; entrance –1; collision 0 (collision parameters are set for each substance at the MS – method parameters); exit 2; LM2 resolution 14.5; HM2 resolution 14.5; Ion energy 25.0 and a multiplier energy of 650 V.

*Multiple reaction monitoring settings for PUFA, eicosanoids/docosanoids semi-quantification*. Method A from 0.0 – 9.0 min for PGF2 349.0 -> 192.7, collision energy 22 eV; PD1 and PD1 isomers like PDX 359,0 -> 153.3, collision energy 17 eV; TXB2 369.0 -> 195.0, collision energy 13 eV, PGE3 349,0 -> 233,0, collision energy 17 eV, LXA4 / LXB4 351,0 -> 115.0, collision energy 12 eV, 8i-PGF3 351.0 -> 193.0, collision energy 22 eV, 20-COOH-LTB4 365.0 -> 195.0, collision energy 13 eV, RvD1, RvD2 375.0 -> 141.3, collision energy 13 eV; RvE1 375.1 -> 141.3, collision energy 13 eV; from 9.0 - 12.5 min for 13-HODE 294.7 -> 170.7, collision energy 16 eV; 9-HODE 294.7 -> 194.7, collision energy 16 eV; 5-HEPE 317.0 -> 115.0, collision energy 17 eV; 12-HEPE 317.0 -> 179.0, collision energy 17 eV; 15-HEPE 317.0 -> 219.0, collision energy 17 eV, LTC4 623.9 -> 272.0, collision energy 14 eV from 12.5 – 16.0 min for LA 279.3 -> 279.0, collision energy 10 eV; 8-HEPE 317.0 -> 255.0, collision energy 17 eV, 18-HEPE 317.0 -> 259.0, collision energy 17 eV; 5-KETE, 12-KETE and 15-KETE 317.0 -> 273.0, collision energy 17 eV, 20-HETE 319.0 > 245.0, collision energy 10 eV; LTC4 623.9 -> 272.0, collision energy 14 eV; from 12.5 – 16.0 min for LA 279.3 -> 59.2, collision energy 25 eV; EPA 301.0 -> 203.2, collision energy 12 eV; AA 303.0 -> 259.3, collision energy 11 eV, DHA 327.1 -> 29.3, collision energy 14 eV.

Method B from 0.0 – 9.8 min for PGE2, d15d12PGD2, PGD2, d15d12PGJ2, PGJ2 315.0 -> 271.3, collision energy 13 eV; LTB5 333.0 -> 195.0, collision energy 13 eV; LTB4 335.0 -> 195.0, collision energy 13 eV; RvE1 349.1 -> 195.3, collision energy 13 eV; HXA3, HXB3, 20-COOH-AA 335.0 -> 273.3, collision energy 13 eV; LXA5 349.0 -> 115.0, collision energy 12 eV; 20-OH-LTB4 351.0 -> 195.0, collision energy 13 eV; MAR 359.0 -> 250.0, collision energy 13 eV; from 9.8 - 12.5 min for 5-HETE 318.7 -> 115.0, collision energy 14 eV; 8-HETE 319.0 -> 155.0, collision energy 14 eV; 11-HETE 319.0 -> 167.0, collision energy 14 eV; 12-HETE 319.0 -> 179.0, collision energy 14eV; 15-HETE 319.0 -> 218.9, collision energy 11 eV; 4-HDHA 343.0 -> 101.0, collision energy 10 eV; 10-HDHA 343.0 -> 181.0, collision energy 10 eV; 14-HDHA 343.0 -> 205.0, collision energy 10 eV; 17-HDHA 343.0 -> 245.0, collision energy 14 eV, 20-HDHA 343.0 -> 285.0, collision energy 10 eV; 13-KODE 293.0 -> 249.0, collision energy 17 eV and from 12.5 – 16.0 min for LA 279.3 -> 59.2, collision energy 25 eV; EPA 301.0 -> 203.2, collision energy 12 eV, AA 303.0 -> 259.3, collision energy 11 eV, DHA 327.1 -> 229.3, collision energy 14 eV.

*Standard solutions*. Stock solutions of the PUFAs, eicosanoids and docosanoids were prepared by dissolving the solutions obtained from Cayman-Chemicals (Tallin, EST), BioMol International (Kastel-Med KFT, Budapest, H), Sigma-Aldrich (Budapest, H), Larodan Lipids (Malmö, S) and Dr. Charles Serhan (Harvard, USA) with methanol to yield a final concentration of 10 μg/mL. All stock solutions were stored in darkness at -80°C. The reference PUFAs, eicosanoids and docosanoids were used for the assay validation.

*Quantification*. Individual eicosanoids and docosanoids were quantified based on the determination of the “Area Under the Curve” (AUC) and compared with the AUC of standard compounds. To ensure optimal extraction isotope labelled standard compounds were used. This analytical procedure was established for liquids and tissue analysis.

*Statistics*. The data are shown as means of triplicate measurements and standard error mean values per data point. One data point represents 5 subjects. Statistical analysis was performed using the program SPSS 16.0 (SPSS Inc., Chicago, IL, USA). A P <0.05 was considered to be significant using student t-test for independent samples.

**Fatty acids analyses**

Plasma samples, thawed in fridge overnight, were vortexed, centrifuged and pipetted into vials. Internal standard (triheptadecanoin) was added and samples were methylated with 3N HCl in Methanol. FAMEs were extracted with hexane, then samples were neutralized with 3N KOH in water. After mixing and centrifuging the hexane phase was injected into the GC-FID

Analysis was performed on a 7890A GC with a split/splitless injector, a 7683B automatic liquid sampler, and flame ionization detection (Agilent Technologies, Palo Alto, CA). Separations were performed on a SP-2380 (30 m × 0.25 mm i.d. × 0.25 µm film thickness) column from Supelco.

**Carotenoid analysis**

25 μL plasma are pipetted into vials and proteins are precipitated and carotenoids extracted with isopropanol added internal standard (β-Apo-8-carotenal). After thorough mixing and subsequent centrifugation, an aliquot of the isopropanol phase is injected into the HPLC-UV. Analysis is performed on a 1100-series HPLC with an 1260 diode array detector (453nm) (Agilent Technologies, Palo Alto, CA ). Separations were performed on a 3 µm, YMC C30 (150 mm × 4.6 mm i.d.) column from YMC (Japan).

***Supplemental Table 1***

**Inclusion criteria**

**Healthy overweight and obese participants, BMI of 25-35 kg/m^2^, age between 50-65 years (inclusive)**

**Exclusion criteria**

- **Weight change of ≥ 3kg in the preceding 3 months**
- **Current smokers**
- **Substance abuse**
- **Excess alcohol intake**
- **Pregnancy**
- **Diabetes**
- **Cardiovascular disease**
- **Cancer**
- **Gastrointestinal disease e.g. inflammatory bowel disease or irritable bowel syndrome**
- **Kidney disease**
- **Liver disease**
- **Pancreatitis**
- **Use of medications likely to interfere with energy metabolism, appetite and hormonal regulation including anti-inflammatory drugs or steroids, antibiotics, androgens, phenytoin, erythromycin and thyroid hormones.**
- **Having metallic or magnetic implants such as pacemakers**
- **Claustrophobia**

***Supplementary table 2***

| **variable** | **subjects** | **EST-EN R** | **subjects** | **CON-EN R** |
| --- | --- | --- | --- | --- |
| alcohol | 65 | 0,75 | 60 | 0,70 |
| red-meat | 62 | 0,79 | 60 | 0,64 |
| meat products | 68 | 0,84 | 63 | 0,45 |
| red meat dishes | 62 | 0,75 | 59 | 0,40 |
| poultry | 64 | 0,82 | 62 | 0,58 |
| poultry dishes | 66 | 0,89 | 60 | 0,66 |
| cheeses | 64 | 0,67 | 63 | 0,34 |
| butter | 66 | 0,73 | 61 | 0,36 |
| whole milk | 63 | 0,68 | 61 | 0,47 |
| milk beverages | 66 | 0,90 | 57 | 0,56 |
| egg | 63 | 0,72 | 61 | 0,62 |
| low fat milk | 65 | 0,75 | 62 | 0,74 |
| yoghurt | 68 | 0,82 | 63 | 0,68 |
| fish | 68 | 0,83 | 62 | 0,61 |
| **animal-based food** | **67** | **0,81** | **63** | **0,60** |
| fruit | 64 | 0,68 | 60 | 0,59 |
| fruit juices | 63 | 0,75 | 62 | 0,48 |
| soups | 63 | 0,76 | 60 | 0,39 |
| vegetables | 63 | 0,80 | 60 | 0,66 |
| **plant-based food** | **62** | **0,80** | **58** | **0,62** |
| biscuits | 60 | 0,81 | 63 | 0,51 |
| confect | 65 | 0,70 | 60 | 0,59 |
| high-energy drinks | 62 | 0,74 | 57 | 0,64 |
| ice creams | 69 | 0,72 | 64 | 0,51 |
| salty snacks | 66 | 0,76 | 61 | 0,59 |
| **snacks/ sweets** | **66** | **0,86** | **61** | **0,69** |
| **general healthy food** | **62** | **0,85** | **58** | **0,51** |
| **general non-healthy food** | **65** | **0,88** | **60** | **0,63** |
| sLUT | 66 | 0,80 | 65 | 0,78 |
| sBCAR | 64 | 0,73 | 63 | 0,74 |
| s9CBC | 65 | 0,88 | 64 | 0,62 |
| sAT-LYC | 63 | 0,85 | 65 | 0,67 |
| **sCARO-sum** | 66 | 0,88 | 64 | 0,66 |
| **s-sum-SAFAs** | 66 | 0,81 | 66 | 0,62 |
| **s-sum-MUFAs** | 66 | 0,73 | 67 | 0,50 |
| **s-sum-n3-PUFAs** | 60 | 0,89 | 62 | 0,72 |
| **s-sum-n6-PUFAs** | **69** | **0,84** | **69** | **0,70** |
| sROL | 68 | 0,78 | 65 | 0,48 |
| sATRA | 64 | 0,78 | 62 | 0,38 |
| s9CDHRA | 65 | 0,87 | 59 | 0,72 |
| **s-sum proinfl. lipid-mediators** | **67** | **0,75** | **61** | **0,41** |
| **s-sum prores. lipid-mediators** | **62** | **0,75** | **57** | **0,64** |
| **RXR-signalling pathway** | **69** | **0,96** | **66** | **0,96** |
| **RAR-signalling pathway** | **71** | **0,96** | **69** | **0,95** |
| **PPAR-signalling pathway** | **71** | **0,91** | **71** | **0,87** |
| **VDR-signalling pathway** | **70** | **0,94** | **72** | **0,91** |
| **NURR1-signalling pathways** | **69** | **0,94** | **68** | **0,90** |
| total-Chol | 65 | 0,76 | 64 | 0,65 |
| HDL-Chol | 67 | 0,80 | 67 | 0,68 |
| LDL-Chol | 66 | 0,86 | 66 | 0,71 |
| sGLUC | 66 | 0,83 | 65 | 0,76 |
| sINS | 68 | 0,82 | 65 | 0,69 |
| sNEFAS | 69 | 0,67 | 68 | 0,61 |
| sTRI | 66 | 0,78 | 65 | 0,58 |
| sVITD | 64 | 0,82 | 64 | 0,70 |
| sVITK2 | 67 | 0,82 | 66 | 0,60 |
| eADIQ | 68 | 0,85 | 67 | 0,70 |
| eIL18 | 66 | 0,85 | 62 | 0,51 |
| BP-dia | 68 | 0,83 | 67 | 0,70 |
| BP-sys | 69 | 0,76 | 64 | 0,63 |
| eosinophils | 65 | 0,85 | 63 | 0,61 |
| monocytes | 67 | 0,70 | 64 | 0,59 |
| HOMA2-IR | 68 | 0,79 | 67 | 0,72 |
|  | 61 | 0,81 | 62 | 0,73 |
| SAT | 65 | 0,77 | 65 | 0,69 |
| IAAT | 61 | 0,82 | 63 | 0,28 |
| IHCL |  |  |  |  |
|  | 67 | 0,87 | 65 | 0,69 |
| indir calorimetry | 64 | 0,69 | 62 | 0,67 |
| RQ |  |  |  |  |
|  | 68 | 0,94 | 69 | 0,97 |
| sex | 68 | 0,77 | 66 | 0,75 |
| BW | 65 | 0,69 | 66 | 0,74 |
| BMI | 66 | 0,80 | 63 | 0,69 |
| HC | 64 | 0,81 | 66 | 0,74 |
| WC | 69 | 0,85 | 68 | 0,65 |
| WHIPR | 64 | 0,78 | 62 | 0,54 |
| HR |  |  |  |  |
|  | 55 | 0,92 | 59 | 0,55 |
| Firmicutes | 57 | 0,68 | 60 | 0,37 |
| Bifidobacterium | 65 | 0,75 | 60 | 0,70 |
| **Mean** |  | **0,80** |  | **0,63** |

**Prediction of variables after intervention using the Elastic-Net model**. Variables were predicted in the “establishment group” (EST) for a transversal prediction using the Elastic-Net (EN) regression models and in addition a “confirmation group” prediction (CON) was performed using the same methodology in the same group of volunteers after intervention using the 72 PBMC transcriptomic data of these volunteers. For each variable the following is reported: the Pearson correlation coefficient (R) between observed and predicted values, and the experimental method used to determine the variables. *For abbreviations check table 1.*

**Supplemental Table 3:.**

| **variable** | **subjects** | **EST-LR R** | **subjects** | **CON-LR R** |
| --- | --- | --- | --- | --- |
| alcohol | 65 | 0,78 | 60 | 0,57 |
| red-meat | 62 | 0,77 | 60 | 0,62 |
| meat products | 68 | 0,79 | 63 | 0,50 |
| red meat dishes | 62 | 0,73 | 59 | 0,32 |
| poultry | 64 | 0,84 | 62 | 0,44 |
| poultry dishes | 66 | 0,81 | 60 | 0,59 |
| cheeses | 64 | 0,63 | 63 | 0,29 |
| butter | 66 | 0,51 | 61 | 0,41 |
| whole milk | 63 | 0,72 | 61 | 0,33 |
| milk beverages | 66 | 0,83 | 57 | 0,55 |
| egg | 63 | 0,75 | 61 | 0,56 |
| low fat milk | 65 | 0,72 | 62 | 0,66 |
| yoghurt | 68 | 0,77 | 63 | 0,60 |
| fish | 68 | 0,77 | 62 | 0,45 |
| **animal-based food** | **67** | **0,74** | **63** | **0,50** |
| fruit | 64 | 0,68 | 60 | 0,39 |
| fruit juices | 63 | 0,72 | 62 | 0,41 |
| soups | 63 | 0,69 | 60 | 0,41 |
| vegetables | 63 | 0,86 | 60 | 0,54 |
| **plant-based food** | **62** | **0,80** | **58** | **0,54** |
| biscuits | 60 | 0,79 | 63 | 0,49 |
| confect | 65 | 0,73 | 60 | 0,58 |
| high-energy drinks | 62 | 0,74 | 57 | 0,55 |
| ice creams | 69 | 0,69 | 64 | 0,51 |
| salty snacks | 66 | 0,72 | 61 | 0,51 |
| **snacks/ sweets** | **66** | **0,81** | **61** | **0,61** |
| **general healthy food** | **62** | **0,86** | **58** | **0,31** |
| **general non-healthy food** | **65** | **0,83** | **60** | **0,59** |
| sLUT | 66 | 0,74 | 65 | 0,65 |
| sBCAR | 64 | 0,74 | 63 | 0,69 |
| s9CBC | 65 | 0,77 | 64 | 0,63 |
| sAT-LYC | 63 | 0,87 | 65 | 0,56 |
| **sCARO-sum** | **66** | **0,81** | **64** | **0,58** |
| **s-sum-SAFAs** | **66** | **0,68** | **66** | **0,63** |
| **s-sum-MUFAs** | **66** | **0,62** | **67** | **0,43** |
| **s-sum-n3-PUFAs** | **60** | **0,83** | **62** | **0,60** |
| **s-sum-n6-PUFAs** | **69** | **0,83** | **69** | **0,63** |
| sROL | 68 | 0,71 | 65 | 0,44 |
| sATRA | 64 | 0,72 | 62 | 0,45 |
| s9CDHRA | 65 | 0,83 | 59 | 0,71 |
| **s-sum proinfl lipid-mediators** | **67** | **0,75** | **61** | **0,20** |
| **s-sum prores lipid-mediators** | **62** | **0,71** | **57** | **0,49** |
| **RXR-signalling pathway** | **69** | **0,96** | **66** | **0,96** |
| **RAR-signalling pathway** | **71** | **0,96** | **69** | **0,95** |
| **PPAR-signalling pathway** | **71** | **0,91** | **71** | **0,85** |
| **VDR-signalling pathway** | **70** | **0,92** | **72** | **0,88** |
| **NURR1-signalling pathways** | **69** | **0,94** | **68** | **0,89** |
| total-Chol | 65 | 0,74 | 64 | 0,59 |
| HDL-Chol | 67 | 0,76 | 67 | 0,63 |
| LDL-Chol | 66 | 0,82 | 66 | 0,63 |
| sGLUC | 66 | 0,76 | 65 | 0,59 |
| sINS | 68 | 0,75 | 65 | 0,63 |
| sNEFAS | 69 | 0,59 | 68 | 0,57 |
| sTRI | 66 | 0,62 | 65 | 0,62 |
| sVITD | 64 | 0,74 | 64 | 0,69 |
| sVITK2 | 67 | 0,82 | 66 | 0,53 |
| eADIQ | 68 | 0,77 | 67 | 0,68 |
| sIL18 | 66 | 0,72 | 62 | 0,56 |
| BP-dia | 68 | 0,76 | 67 | 0,67 |
| BP-sys | 69 | 0,76 | 64 | 0,53 |
| eosinophils | 65 | 0,82 | 63 | 0,45 |
| monocytes | 67 | 0,68 | 64 | 0,47 |
| HOMA2-IR | 68 | 0,77 | 67 | 0,64 |
|  | 61 | 0,78 | 62 | 0,69 |
| SAT | 65 | 0,76 | 65 | 0,70 |
| IAAT | 61 | 0,83 | 63 | 0,15 |
| IHCL |  |  |  |  |
|  | 67 | 0,91 | 65 | 0,64 |
| indir calorimetry | 64 | 0,67 | 62 | 0,44 |
| RQ |  |  |  |  |
|  | 68 | 0,93 | 69 | 0,98 |
| sex | 68 | 0,74 | 66 | 0,73 |
| BW | 65 | 0,66 | 66 | 0,61 |
| BMI | 66 | 0,76 | 63 | 0,53 |
| HC | 64 | 0,78 | 66 | 0,71 |
| WC | 69 | 0,81 | 68 | 0,58 |
| WHIPR | 64 | 0,77 | 62 | 0,52 |
| HR |  |  |  |  |
|  | 55 | 0,89 | 59 | 0,49 |
| Firmicutes | 57 | 0,51 | 60 | 0,39 |
| Bifidobacterium | 65 | 0,78 | 60 | 0,57 |
| **Mean** |  | **0,77** |  | **0,57** |

**Prediction of variables after intervention** **using the Linear-Regression model.** Variables were predicted in the “establishment group” (EST) for a transversal prediction using the Linear-Regression (LR) model and in addition a “confirmation group” prediction (CON) was performed using the same methodology in the same group of volunteers after intervention using the 72 PBMC transcriptomic data of these volunteers. For each variable the following is reported: the Pearson correlation coefficient between observed and predicted values, and the experimental method used to determine the variables. *For abbreviations check table 1.*
